# Oxymetazoline for the treatment of rosacea-associated persistent facial erythema. Systematic review and meta-analysis protocol

**DOI:** 10.1101/2020.05.25.20111344

**Authors:** Geovanna Cárdenas, Francisco Novillo, Shuheng Lai, Héctor Fuenzalida, Francisca Verdugo, Gabriel Rada

## Abstract

**Objective:** The objective of this systematic review is to assess the impact of oxymetazoline in patients with moderate to severe rosacea.

**Data Sources:** We will conduct a comprehensive search in PubMed/Medline, Embase, Cochrane Central Register of Controlled Trials (CENTRAL), Lilacs, the International Clinical Trials Registry Platform (ICTRP), ClinicalTrials.gov, US National Institutes of Health (NIH) and grey literature, to identify all relevant randomized controlled trials regardless of language or publication status (published, unpublished, in press and in progress).

**Eligibility criteria for selecting studies and methods:** We will include randomized trials evaluating the effect of oxymetazoline in patients with moderate to severe rosacea. Two reviewers will independently screen each study for eligibility, data extraction, and assess the risk of bias. We will pool the results using meta-analysis and will apply the GRADE [1] system to assess the certainty of the evidence for each outcome.

**Ethics and Dissemination:** No ethics approval is considered necessary. The results of this review will be widely disseminated via peer-reviewed publications, social networks and traditional media.

**Protocol and Registration:** This protocol was adapted to the specificities of the question assessed in this review and registered to PROSPERO with the ID CRD42020150262.

## INTRODUCTION

Rosacea is a chronic inflammatory skin disease that is characterized by persistent facial erythema, papules, pustules, telangiectasia and recurrent flushing; affecting primarily the cheeks, nose, chin, and forehead [2] [3] [4]. Rosacea affects both women and men equally, with a higher rate amongst people between ages 45 and 60 [5]. Recent findings suggest rosacea might appear also in people of different ethnicity and darker phototype [6].

The pathogenesis of rosacea remains unclear, but genetic factors, overgrowth of commensal organisms, the dysregulation of innate immunity and of the neurovascular system may be involved [7]. Rosacea is characterized by repeated remissions and exacerbations, which results from well described risk factors or triggers, divided into exogenous (ultraviolet radiation exposure, heat, spicy food, stress, alcohol, some medications) and endogenous (hormonal changes, body mass index) [2] [8] [9].

In general, the recommendations for the initial treatment of rosacea includes avoidance of trigger factors, usage of sunscreen, application of moisturizers and gentle cleansing of the whole face. [7] [10]. In the case of persistent facial erythema previously included in the erythematotelangiectatic type of rosacea, brimonidine tartrate, a topical selective α-adrenergic receptor agonist with vasoconstrictive activity has been approved for treatment of persistent facial erythema [7].

Recent clinical trials have been studying the safety and efficacy of oxymetazoline hydrochloride as a new topical to treat persistent erythema in adults. Oxymetazoline is a selective α-adrenoreceptor agonist, and vasoconstrictor of the cutaneous microvasculature [11] [12]. Topical application of oxymetazoline will reduce erythema associated with rosacea through vasoconstrictive effects on the α1-adrenoreceptors [13].

The aim of this systematic review is to provide a rigorous up to date summary of the available evidence on the role of oxymetazoline in the treatment of patients with rosacea-associated persistent facial erythema.

## METHODS

### Protocol and Registration

This manuscript complies with the Preferred Reporting Items for Systematic review and Meta-Analysis for reporting systematic reviews and meta-analyses.

This protocol was adapted to the specificities of the question assessed in this review and registered to PROSPERO with the ID CRD42020150262.

### Eligibility Criteria

#### Types of studies

We will include randomized controlled trials (RCTs) meeting our inclusion criteria and reporting useable data. Quasi-randomized trials (QRTs) will not be included. We will exclude studies evaluating the effects on animal models or in vitro conditions.

#### Types of participants

We will include trials assessing participants aged 18 and above with a diagnosis of moderate to severe erythematotelangiectatic rosacea, defined as ≥grade 3 on the Clinician Erythema Assesment (CEA) and the Subject Self-Assessment for rosacea facial redness (SSA). Participants with another dermatologic condition being treated, systemic diseases and hypersensitivity to oxymetazoline will be excluded.

#### Types of interventions

We will include trials evaluating the impact of topical application of oxymetazoline for the treatment of rosacea-associated persistent facial erythema. The comparison of interest will be placebo (oxymetazoline hydrochloride cream versus placebo). We will not restrict our criteria to any dosage, duration or timing of administration.

#### Types of outcome measures

We will not use outcomes as an exclusion criteria during the selection process. Any article meeting all the criteria except for the outcome criterion will be preliminarily included and assessed in full text. We will consider the following outcomes:

Primary outcomes
- Clinician Erythema Assessment (CEA) and the Subject Self-Assessment for rosacea facial redness (SSA)

Secondary outcomes
- Rebound of erythema

Other outcomes
- Total adverse events

### Search strategies

#### Electronic searches

We will conduct a comprehensive search in PubMed/Medline, Embase, Cochrane Central Register of Controlled Trials (CENTRAL), Lilacs, the International Clinical Trials Registry Platform (ICTRP), ClinicalTrials.gov, US National Institutes of Health (NIH) and grey literature, to identify all relevant randomized controlled trials regardless of language or publication status (published, unpublished, in press and in progress). The searches will cover from the inception date of each database until the day before submission.

The following strategy will be used to search in Pubmed/Medline Database. We will adapt it to the syntax of other databases.

1 Rosacea[Mesh] 2 erythematotelangiectatic 3 rosacea* 4 rhinophyma* 5 (pyoderma AND faciale*) 6 1 OR 2 OR 3 OR 4 OR 5 7 "Oxymetazoline"[Mesh] 8 oxymetazolin* 9 Otrivin 10 Afrin 11 Operil 12

Dristan 13 Dimetapp 14 Oxyspray 15 Facimin 16 Nasivin 17 Nostrilla 18 Utabon 19 Sudafed OM 20 Vicks Sinex 21 Zicam 22 SinuFrin 23 Drixoral 24 Mucinex Full Force 25 7 OR 8 OR 9 OR 10 OR 11 OR 12 OR 13 OR 14 OR 15 OR 16 OR 17 OR 18 OR 19 OR 20 OR 21 OR 22 OR 23 OR 24 26 randomized controlled trial [pt] 27 controlled clinical trial [pt] 28 randomized [tiab] 29 placebo [tiab] 30 drug therapy [sh] 31 randomly [tiab] 32 trial [tiab] 33 groups [tiab] 34 25 OR 26 OR 27 OR 28 OR 29 OR 30 OR 31 OR 32 OR 33 35 animals [mh] NOT humans [mh] 36 34 NOT 35 37 6 AND 25 AND 36.

#### Searching other resources

In order to identify articles that might have been missed in the electronic searches, we will do the following:

1. MEDLINE for systematic reviews addressing the same question as our review.
2. In order to find additional literature we will run a Google Scholar search for key terms and authors.
3. We will handsearch reference lists of all included studies and of relevant reviews retrieved by the electronic searching to identify further relevant trial.
4. Authors of included studies will be contacted for any additional published or unpublished data.

#### Selection of studies

The results of the literature search will be uploaded to the screening software Collaboratron^™^[14].

In Collaboratron^™^ [14] two researchers will independently screen the titles and abstracts yielded by the search against the inclusion criteria. We will obtain full-text reports for all potentially eligible studies that appear to meet the inclusion criteria or require further evaluation to decide about their inclusion. The same review authors will assess independently those articles and decide on fulfilment of our inclusion criteria. A third review author will resolve discrepancies in the case of disagreement. Articles retrieved from the screening and included in the review will be recorded in RevMan 5.3 [15]. Excluded trials after full text revision and the primary reason for the decision will be listed.

The selection process will be documented in a in a PRISMA [16] flow diagram adapted for the purpose of this project.

#### Data extraction and management

Using standardized forms, two reviewers will independently extract data from each included study. We will collect the following information: study design, setting, baseline characteristics of patients and study eligibility criteria, details of intervention including dose and duration, number of patients assigned to each group, the outcomes assessed and the time they were measured; number of patients with adverse reactions per treatment group and method used to seek adverse reactions. Losses to follow up, exclusions, and the reasons accordingly. A third reviewer will resolve discrepancies in the case of disagreement.

#### Risk of bias assessment

Risk of bias in included studies will be assessed according to the ‘Risk of bias’ table, which is the tool recommended by The Cochrane Collaboration [17]. Descriptions and judgements about the following domains for each study will be included: adequacy of sequence generation, allocation concealment, blinding, addressing of incomplete outcome data, likelihood of selective outcome reporting, and other potential sources of bias.

The ‘Risk of bias’ table will be prepared independently by two review authors, with a third review author acting as arbiter. Original study authors will be contacted for further information if one of the review authors requires clarification wherever necessary.

#### Measures of treatment effect

We will report dichotomous outcomes as risk ratio (RR) or odds ratio (OR), with 95% confidence interval (CI) and the continuous outcomes will be reported as mean difference (MD) with 95% CI. Outcomes using different scales will be reported as standardized mean differences (SMD).

Then, these results will be displayed on the ‘Summary of Findings Table’ as mean difference.

#### Dealing with missing data

We will contact the main authors of trials with missing data in order to verify key study characteristics and obtain missing numerical outcome data. If we don’t receive any reply, we will analyze the available data. The potential impact of the “missing data” will be addressed in the Discussion section using the analysis of the best and the worst scenario.

If no intention to treat (ITT) analysis has been reported or and modified ITT it’s been reported, we will reanalyze the data following intention to treat principles if possible.

#### Assessment of Heterogeneity

Heterogeneity will be quantitatively assessed with a statistical test (Q statistic) and the I2 statistic. Statistically significant heterogeneity will be defined as at least one positive test (establishing a cut-off value of P = 0.10 for the Mantel-Haenszel Chi2 test, or values over 50% using the I2 statistic)

#### Assessment of Reporting Biases

We will visually assess the presence of publication bias with the use of funnel plots. Evidence of asymmetry will be based on P < 0.10, and present intercepts with 90% CIs. Other reporting biases, including outcome reporting bias, will be evaluated through discrepancies between the registered protocol and the final publication. If we cannot find the record of a study in the WHO International Clinical Trials Registry Platform authors will be contacted for further information.

#### Data synthesis

Statistical analysis will be performed in accordance with the guidelines for statistical analysis developed by Cochrane [13]. If possible, all studies will be pooled to perform meta-analysis comparing any topical use of oxymetazoline vs placebo for each outcome measure.

When possible, meta-analyses using the random-effects inverse variance model will be carried out to estimate the pooled measure of treatment effect, fixed effects models will be performed to evaluate if findings are not sensitive to choice of analysis. If specific populations or interventions are found to be significantly different, separate meta-analyses will be performed to assess if heterogeneity is explained by some of these, or if a convincing subgroup effect is found. Discrepancies found between random and fixed effects analyses, will be addresses in in the ‘Discussion’.

#### Subgroup analysis and investigation of heterogeneity

If enough data is available, subgroup analysis will be performed to assess different presentation and concentration of oxymetazoline.

#### Sensitivity analysis

Sensitivity analyses will be performed to evaluate the impact of the inclusion or exclusion of missing data and the choice of a fixed-effect or random-effects models.

#### Summary of findings

The certainty of the evidence for all outcomes will be reviewed using the Grading of Recommendations Assessment, Development and Evaluation working group methodology (GRADE Working Group) [1]. Findings for the main outcomes will be summarized in Summary of Findings (SoF) tables.

## Data Availability

All data related to the project will be available on the final publication.

## NOTES

### Roles and Contributions

GC conceived the protocol. GC, SL, FN y HF drafted the manuscript, and all other authors contributed to it. The corresponding author is the guarantor and declares that all authors meet authorship criteria and that no other authors meeting the criteria have been omitted.

### Ethics

As researchers will not access information that could lead to the identification of an individual participant, obtaining ethical approval was waived.

### PROSPERO Registration

This protocol has been submitted PROSPERO Registration ID: CRD42020150262.

